# Effectiveness of the EPA-based ‘Toolbox Family Medicine’ on students’ learning satisfaction: study protocol for a controlled trial

**DOI:** 10.1101/2022.01.18.22269060

**Authors:** Nicola Amarell, Maximilian Wehner, Johanna Schroeder, Dorothea Wild, Thomas Welchowski, Birgitta Weltermann

## Abstract

**Background:** Family practices constitute an important learning environment for medical students. However, teaching situations markedly vary between practices, and students frequently find rotations underwhelming. Especially, students’ active participation in patient care varies profoundly, although it has a significant impact on students’ interest in primary care careers. To standardize and improve learning situations in practices, we developed the so-called ‘Toolbox Family Medicine (TFM)’ using the concept of entrustable professional activities. It provides standardized learning content appropriate for students’ levels and allows teaching adaptable to actual practice conditions.

**Methods:** Using a controlled trial with a waiting list control arm, we will evaluate the effectiveness of the toolbox on students’ learning satisfaction. A total of 94 students will be allocated 1:1 to intervention and control practices. The teaching concept ‘Toolbox Family Medicine (TFM)’ comprises a didactic workshop for supervising physicians and a toolbox with practice-specific tasks for medical students. The primary outcome is students’ overall satisfaction with their learning progress after the rotation. Secondary outcomes include the kind and number of tasks performed, the entrusted level per task, the feasibility of implementing the toolbox in actual practice settings, and students’ motivation to pursue a career in primary care.

**Discussion:** We assume an improvement in learning satisfaction with the intervention. The study will begin with the next practice rotations.

**Administrative information:** 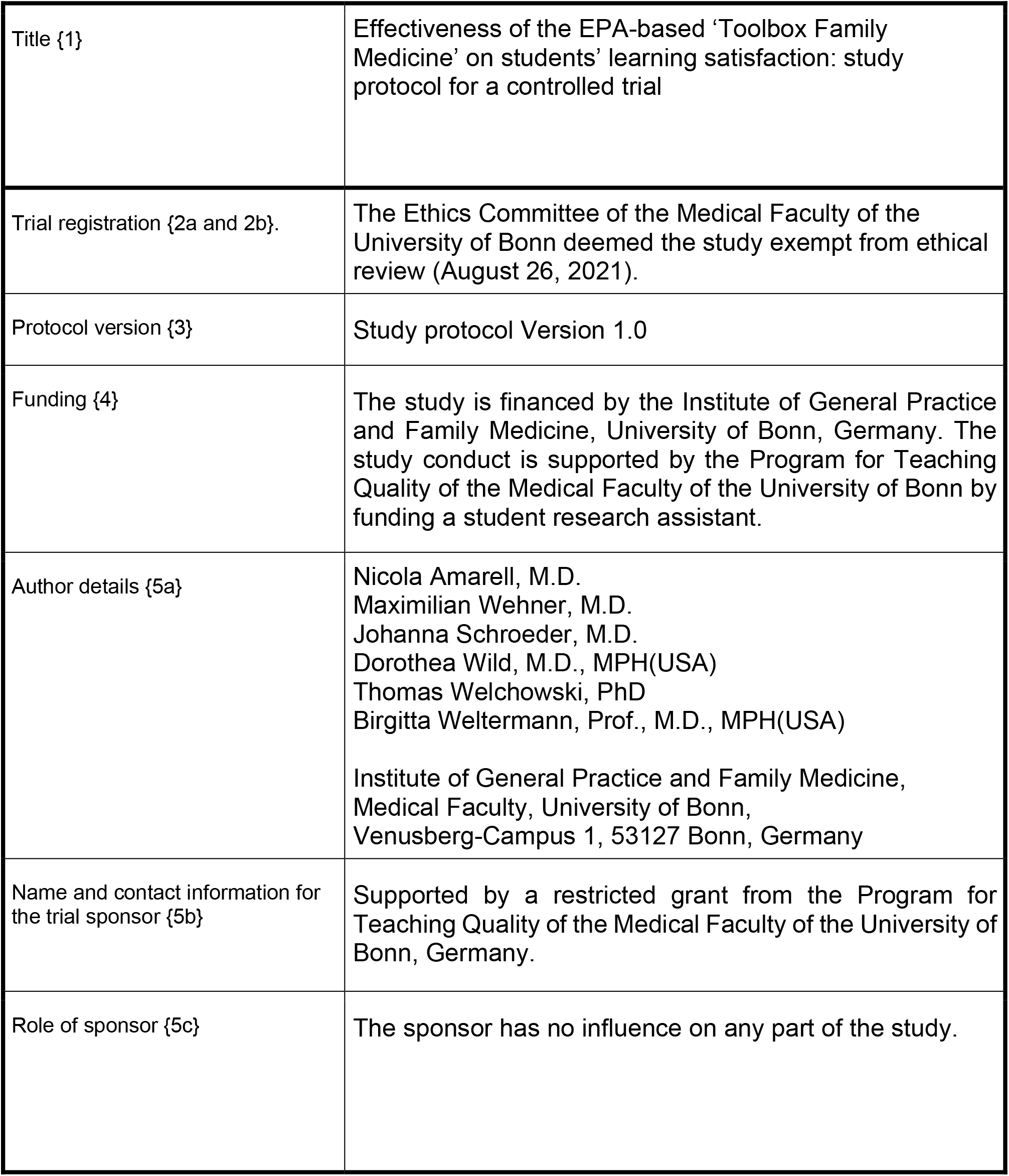

## Background

### Background and rationale {6a}

Family practices constitute an important learning environment for medical students (Pershing et al., 2013). However, practice rotations are heterogeneous teaching situations that students often find underwhelming (Herwig et al., 2017). The extent of students’ participation varies considerably, although it significantly affects students’ interest in primary care careers (Herwig et al., 2017). There are several reasons for unsatisfactory practice rotations: Supervising physicians balance competing tasks as they provide teaching integrated into their daily patient services. In addition, time pressure and lack of didactic training impair teaching quality.

Aiming at a better and more standardized learning environment in family practices, we developed the so-called ‘Toolbox Family Medicine’ (TFM). It provides standardized learning content suitable for students’ levels and allows teaching adaptable to actual practice conditions. Learning content is structured following the concept of entrustable professional activities (EPA). All EPAs describe small tasks typical for routine primary care that students have to perform guided by the respective teaching physician (Ten Cate et al., 2015). Completing an EPA, students receive feedback from the supervisor on their performance, which provides the basis for future entrustment decisions. This linkage of activity and supervision level creates a performance-based training, so-called competency-based medical education. The overall aim is to gradually reduce supervision and increase students’ responsibilities based on individual progress (Peters et al., 2017). The five grades described by Chen (2015) are “allowed to observe” (grade 1), “direct supervision” (grade 2), “indirect supervision” (grade 3), “acting unsupervised” (grade 4), “guiding others” (Grade 5) (see Table 1).

**Table 1:** EPA supervision levels (Chen et al. 2015)

|  |  |
| --- | --- |
| Level 1 | Not allowed to practice EPA<br>a) Inadequate knowledge/ skill; not allowed to observe<br>b) Adequate knowledge/ some skill, allowed to observe |
| Level 2 | May act under proactive, ongoing, full supervision (direct)<br>a) As coactivity with supervisor<br>b) With supervisor in room, ready to step in as needed |
| Level 3 | May act under reactive supervision (indirect)<br>a) With supervisor immediately available, all findings double-checked<br>b) With supervisor immediately available, key findings double-checked<br>c) With supervisor distantly available, findings reviewed |
| Level 4 | May act unsupervised |
| Level 5 | Allowed to supervise others in the practice of EPA |

Starting in Canada, this concept of competency-based medical education has already been implemented in several countries as a didactic approach for the graduate level. In Germany, EPAs have just started to become part of medical undergraduate and graduate education. Academic teachers of the Charité Berlin University defined twelve core EPAs, grouped into 5 EPA domains, to measure preparedness for entry into residency: 1. along the clinical encounter, 2. general medical procedures, 3. communicating with patients, 4. communicating and collaborating with colleagues, 5. patient care in special situations (Holzhausen et al., 2019). A pilot study described the implementation of EPAs with 62 final year students in four German universities concluding that more work is needed to integrate EPAs in medical curricula (Schick et al., 2019).

Our TFM is an approach to implement EPAs in general practice rotations of undergraduate students. Students and supervisors will be surveyed directly after the rotation. Students’ overall satisfaction with their learning progress is the primary outcome of this controlled trial.

### Objectives {7}

This study evaluates the effectiveness of the teaching concept TFM on students’ overall satisfaction with their learning progress after the rotation. The evaluation will compare students from practices with TFM use (intervention) and without (control). We hypothesize that the TFM will improve learning satisfaction compared to traditional training.

### Trial design {8}

This teaching quality improvement project will be performed as an intervention trial. Teaching practices and students will be allocated in a groupwise manner. The first 47 students will serve as the control group. The next 47 students will serve as the intervention group using the toolbox.

This approach is needed to avoid contamination on the student or practice level, i.e. a practice will not educate control group students after participating in the toolbox teaching. All control practices will receive the toolbox education and materials after the study’s completion (waiting list control approach).

## Methods: Participants, interventions and outcomes

### Study setting {9}

The study will be conducted in family practices accredited by the Institute of General Practice and Family Medicine, University of Bonn. They are located in the greater Bonn area. As part of the undergraduate curriculum, medical students have a two-week practice rotation in family medicine, which takes place in their last clinical semester prior to the final year. All practices and students of the next two terms will be included in the quality improvement project.

### Eligibility criteria {10}

Inclusion and exclusion criteria for practices: Family practices are eligible if they are accredited teaching practices of the Institute of General Practice and Family Medicine, University of Bonn.

Inclusion and exclusion criteria for students: Participating students must be enrolled at the University of Bonn and scheduled for the family practice rotation. Students will be excluded when changing the practice during their rotation.

### Who will take informed consent? {26a}

The Ethics Committee of the Medical Faculty of the University of Bonn has agreed to the teaching quality improvement project and deemed it exempt from review on August 26, 2021. Teaching practices and students participate on a voluntary basis.

### Additional consent provisions for collection and use of participant data and biological specimens {26b}

Not applicable, because no biological specimen will be obtained.

## Interventions

### Explanation for the choice of comparators {6b}

Students of the control group will complete the usual general practice rotation in practices without TFM intervention.

### Intervention description {11a}

The intervention ‘Toolbox Family Medicine’ comprises (1) a TFM training for teaching physicians focused on didactics, (2) the Toolbox Family Medicine with the EPAs and (3) an EPA-compendium, recording the performed EPAs with the respective supervision level (see Table 1).

The TFM intervention will be applied in a two-week rotation in intervention practices.

Elements of the ‘Toolbox Family Medicine’ (TFM):

1. TFM training for teaching physicians:
  a. Invitation email with a brief explanation of the new concept.
  b. Practice visits to hand over and introduce the TFM card box.
  c. One-hour online workshops for supervising physicians: Groups of six to ten teaching physicians will be educated on the concept and use of the toolbox focusing on didactics (e.g. feedback and teachable moments).
2. Toolbox with EPAs (index card box):
  a. Index cards with information on EPAs, feedback rules, application of the toolbox.
  b. Index cards with short and more complex tasks supplemented by the recommended supervision levels, more in-depth information and questions.

Following the supervision levels for medical education (see Table 1) (Chen et al., 2015), we defined limits for the performance of the various medical activities for undergraduate medical education (see Table 2). During their two-week rotation, students may perform EPAs up to a level of 3a.

**Table 2:** List of 72 EPAs and targeted supervision level for rotations in family practices; Core-EPAs are marked with a grey background.

| # | EPAs | Supervision level for two-week rotation |
| --- | --- | --- |
| **1 Medical history** |  |  |
| 1.1 | Past medical history | 1b-3a |
| 1.2 | Medication | 1b-3a |
| 1.3 | Risk factors | 1b-3a |
| 1.4 | Review of systems | 1b-3a |
| 1.5 | Mental and psychological history | 1b-3a |
| 1.6 | Social history | 1b-3a |
| 1.7 | Family history | 1b-3a |
| 1.8 | Occupational history | 1b-3a |
| **2 Physical examination** |  |  |
| 2.1 | Head (incl. cranial nerves) | 1b-3a |
| 2.2 | Neck | 1b-3a |
| 2.3 | Chest | 1b-3a |
| 2.4 | Abdomen | 1b-3a |
| 2.5 | Skin | 1b-3a |
| 2.6 | Musculoskeletal system | 1b-3a |
| 2.7 | Mental status and nervous system | 1b-3a |
| 2.8 | Vascular status | 1b-3a |
| **3 Skills** |  |  |
| 3.1 | Blood pressure measurement | 1b-3a |
| 3.2 | Urine test (dip stick) | 1b-3a |
| 3.3 | Venous blood draw | 1b-3a |
| 3.4 | Electrocardiogram | 1b-3a |
| 3.5 | Measurement of blood glucose (finger stick) | 1b-3a |
| 3.6 | Ankle-Brachial-Index | 1b-3a |
| 3.7 | Geriatric assessment | 1b-3a |
| 3.8 | Subcutaneous/ intramuscular injection (e.g. vaccination) | 1b-2b |
| 3.9 | Spirometry | 1b-3a |
| 3.10 | Otoscopy | 1b-3a |
| 3.11 | Wound care | 1b-2b |
| 3.12 | Abdominal ultrasound | 1b-2b |
| 3.13 | Thyroid ultrasound | 1b-2b |
| 3.14 | Exercise ECG | 1b-2b |
| 3.15 | Interpretation of laboratory findings | 1b-3a |
| 3.16 | Medication adjustment for renal dysfunction | 1b-3a |
| 3.17 | Medication adjustment in geriatric patient | 1b-3a |
| **4 Communication** |  |  |
| 4.1 | Advice on weight reduction | 1b-3a |
| 4.2 | Importance of long-term blood pressure therapy | 1b-3a |
| 4.3 | Alcohol cessation | 1b-2b |
| 4.4 | Smoking cessation | 1b-2b |
| 4.5 | Illness theory and therapy adherence | 1b-3a |
| 4.6 | Consultation on need and conduct of vaccinations | 1b-2b |
| 4.7 | Intercultural aspects of medical care | 1b-3a |
| 4.8 | Living will/ health care proxy | 1b-3a |
| 4.9 | Osteoporosis | 1b-3a |
| **5 Chief complaints** |  |  |
| 5.1 | General check-up, 20-30 year-old patient | 1b-3a |
| 5.2 | General check-up, 50-60 year-old patient | 1b-3a |
| 5.3 | General check-up, > 75 year-old patient | 1b-3a |
| 5.4 | Home visit | 1b-2b |
| 5.5 | Nursing home visit | 1b-2b |
| 5.6 | Dysuria | 1b-3a |
| 5.7 | Headache | 1b-3a |
| 5.8 | Chest pain | 1b-3a |
| 5.9 | Low back pain | 1b-3a |
| 5.10 | Hip pain | 1b-3a |
| 5.11 | Knee pain | 1b-3a |
| 5.12 | Shoulder pain | 1b-3a |
| 5.13 | Neck pain | 1b-3a |
| 5.14 | Upper abdominal pain | 1b-3a |
| 5.15 | Lower abdominal pain | 1b-3a |
| 5.16 | Diarrhea | 1b-3a |
| 5.17 | Sleep disorders | 1b-3a |
| 5.18 | Dizziness | 1b-3a |
| 5.19 | Acute psycho-social stress | 1b-2b |
| 5.20 | Vaccination advice (incl. Vaccination) | 1b-2b |
| 5.21 | Disease management program (DMP) type 2 diabetes | 1b-3a |
| 5.22 | DMP asthma | 1b-3a |
| 5.23 | DMP coronary heart disease | 1b-3a |
| 5.24 | DMP heart failure | 1b-3a |
| 5.25 | Cough | 1b-3a |
| 5.26 | Joint pain | 1b-3a |
| 5.27 | Sore throat and ear ache | 1b-3a |
| 5.28 | Preoperative assessment | 1b-2b |
| 5.29 | Post-discharge visit | 1b-2b |
| 5.30 | Emergency situation management (focus on handover to emergency medical services) | 1b-2b |

An expert panel of five experienced supervising physicians and three GPs in training defined a total of 72 EPAs in five fields: medical history (1), physical examination (2), skills (3), communication (4), chief complaints (5). For details see Table 2.

For the two-week, full-day rotation in family practices, 21 EPAs were selected as core EPAs obligatory for all students. The teaching physician may select additional EPAs. After performing an EPA, the teaching physician will provide feedback to the student on the entrusted level. Additional information and questions on the index cards provide an orientation for the teaching physician for more in-depth teaching related to the EPA. Students should reach an entrustment level of 2b or 3a on all core EPAs, as defined in Table 2. Hence, respective EPAs may be performed and supervised several times until the student reaches the required entrustment level (Peters et al., 2017).

### Criteria for discontinuing or modifying allocated interventions {11b}

Teaching physicians who are invited to the intervention but unable to attend the scheduled TFM training are assigned to the control group. The statistical analyses will account for this.

### Strategies to improve adherence to interventions {11c}

Practices and students are routinely provided with the contact information of the teaching secretary (phone number, email) to report any problem prior or during the rotation.

### Relevant concomitant care permitted or prohibited during the trial {11d}

Not applicable as the study is a teaching intervention.

### Provisions for post-trial care {30}

Not applicable as the study is a teaching intervention.

### Outcomes {12}

#### Primary outcome

The primary outcome is the students’ overall satisfaction with their learning progress using a five-point Likert scale.

#### Secondary outcomes

The student rotation in family practices has been routinely evaluated for years. This evaluation will continue with additional questions regarding satisfaction with their practice placement using the Placement Evaluation Tool (PET), which was initially created for nursing placements in Australia (Cooper et al., 2020) and translated to German (Schroeder et al., 2021). Students of the control and intervention groups will receive the same survey.

Teaching physicians of both groups will receive questions on the execution of the two-week rotation and questions on students’ performance, the latter of which are matched to the PET questionnaire. In intervention practices only, the following data will be obtained: satisfaction with the toolbox content, practicality of the index card box approach, feasibility of implementing the concept in everyday practice flow; students’ record of the performed EPAs and the supervision levels entrusted by the teaching physician.

### Participant timeline {13}

The timeline for teaching physicians and students is outlined in Figure 1.

**Figure 1:**
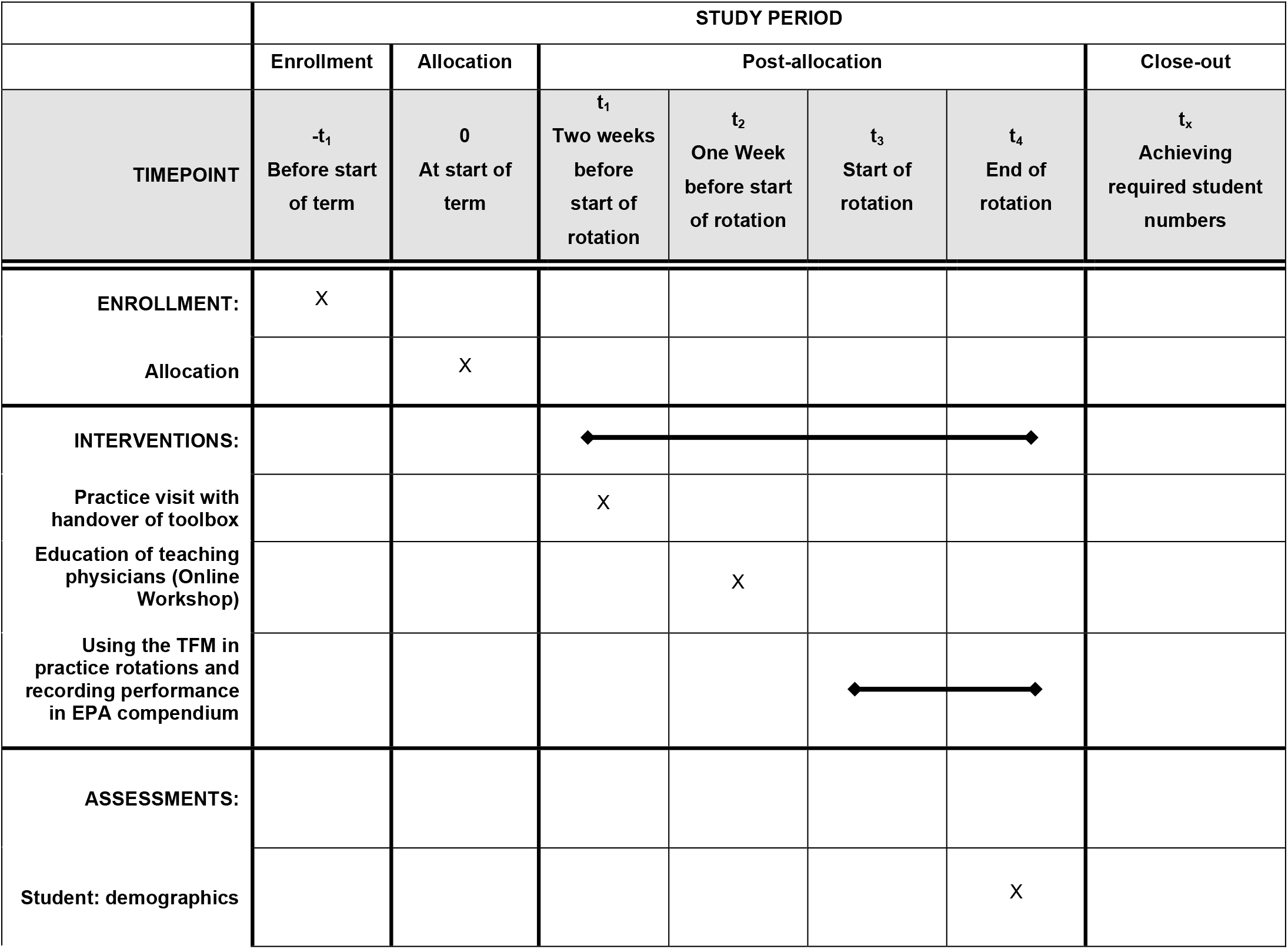
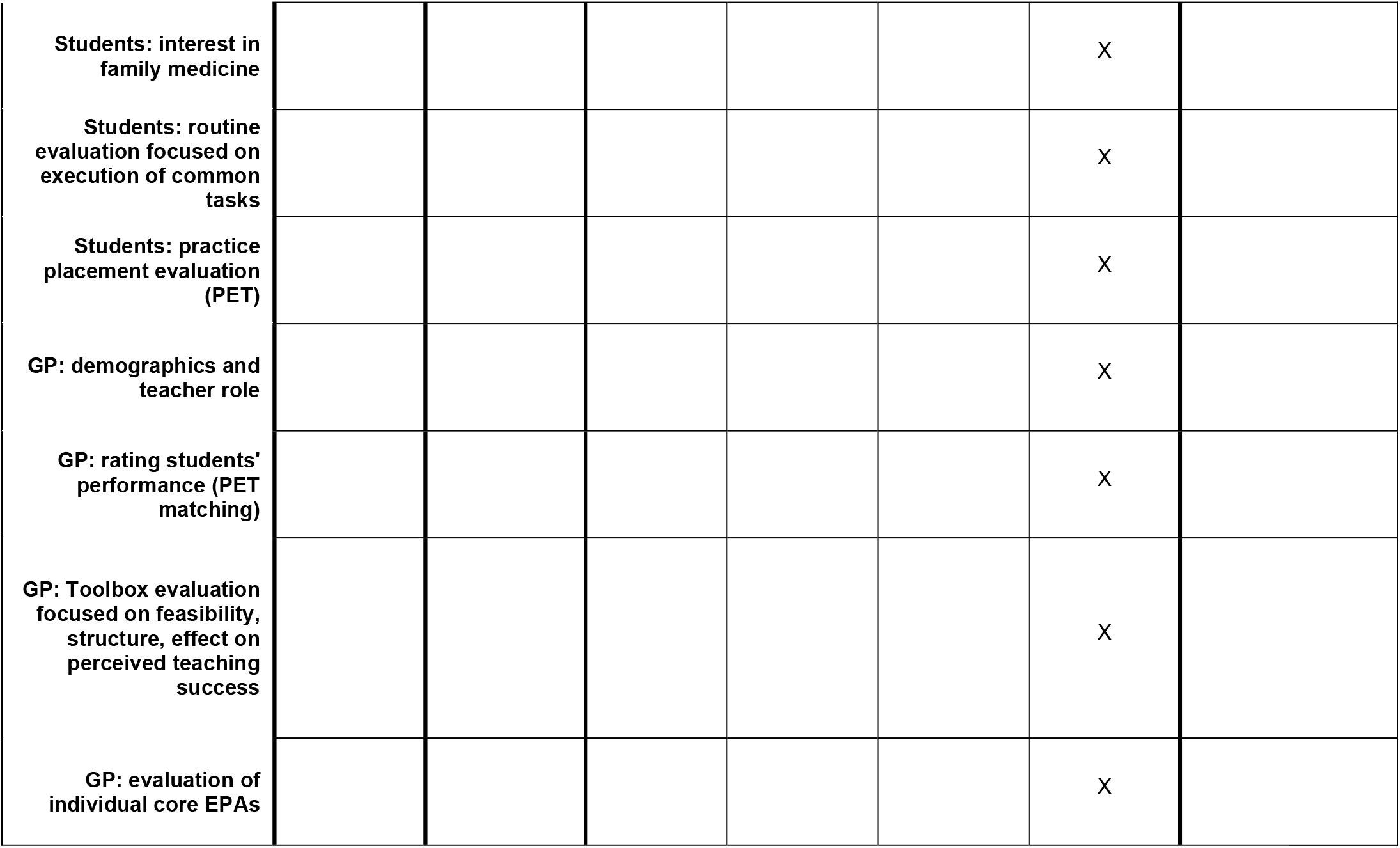
Schedule of enrolment, interventions, and assessments.

### Sample size {14}

The sample size was calculated based on the primary outcome (students’ overall learning satisfaction). The primary outcome will be measured according to German university grades ranging from 1 (best) to 5 (worst). The null hypothesis is that the median category of the primary outcome is equal between control and intervention groups. The Wilcoxon-Mann-Whitney-Test was used to determine the sample size based on the approach of Happ (2019). This method requires prior information about the control and intervention groups, which was retrieved from the study of Herwig et. al. (2017). For the control group, we assumed the absolute frequencies of the group “interest in primary care decreased” for the overall rating of the clerkship as prior information. The term “prior difference” is defined as difference between absolute frequencies of the primary outcome in the “increased” and “decreased” groups in overall rating of clerkship. We assumed half of the prior difference as a minimally relevant educational effect that we are interested in detecting. Based on this assumption, the prior sample of the intervention group was created by adding half of the prior difference to the prior sample of the control group. Furthermore, a significance threshold alpha of 5 %, power level of 80 %, and a student dropout of 5 % is applied to determine the sample size. As a result, this study aims for a controlled trial with at least 94 medical students (47 intervention, 47 control).

### Recruitment {15}

Scheduling for rotations will take place as usual. Practice rotations are part of the compulsory medical school curriculum. Family practices are invited by email and by phone. A complete timeline is presented in Figure 1.

## Assignment of interventions: allocation

### Sequence generation {16a}

Using the routine academic schedule for upcoming rotations in family practices, half of the students and respective practices will serve as the control group. The second half of the students and respective practices will serve as the intervention group. Teaching physicians of the intervention group will be trained successively in small groups before new students are assigned to their practices. Practices participate based on availability.

### {16b}

The allocation sequence is based on the academic year schedule for practices and students. No additional allocation mechanisms are needed.

### Implementation {16c}

Enrollment and assignment of students to practices will be performed by the teaching office as usual. After reaching the planned number of control practices, the subsequent practices will serve as intervention practices.

## Assignment of interventions: Blinding

### Who will be blinded {17a}

No blinding. Students and physicians of the intervention group are aware of the toolbox usage.

### Procedure for unblinding if needed {17b}

Not applicable.

## Data collection and management

### Plans for assessment and collection of outcomes {18a}

Finishing their two-week rotation students will be invited to complete a survey on eCampus, the teaching platform of the University of Bonn. Teaching physicians will be invited to evaluate each rotation using a professional survey platform following data protection laws.

### Plans to promote participant retention and complete follow-up {18b}

Physicians and students will receive up to two reminders to promote follow-up.

### Data management {19}

#### Confidentiality {27}

All data will be stored following standards for data protection and data security at the Institute for General Practice and Family Medicine

University of Bonn, University Hospital Bonn

Venusberg-Campus 1

53127 Bonn, Germany.

All person-level data will be stored in an access-restricted master file. Data analyses will be performed with pseudonymized data only.

### Plans for collection, laboratory evaluation and storage of biological specimens for genetic or molecular analysis in this trial/future use {33}

Not applicable. No samples collected.

## Statistical methods

### Statistical methods for primary and secondary outcomes {20a}

This protocol contains the entire statistical plan. Data management will follow standardized procedures (SOPs, Standard Operating Procedures). Primary and secondary outcomes will be analyzed using descriptive statistics. Inferential statistics will be performed for the primary outcome and selected secondary outcomes to analyze for predictors of learning satisfaction.

### Interim analyses {21b}

No interim analysis is planned.

### Methods for additional analyses (e.g. subgroup analyses) {20b}

Subgroup analysis will be performed by practice clusters.

### Methods in analysis to handle protocol non-adherence and any statistical methods to handle missing data {20c}

Data imputation with standard procedures will be performed if needed to handle missing data (Van Buuren S, 2018).

### Plans to give access to the full protocol, participant level-data and statistical code {31c}

The datasets generated and analysed during the study will not publicly available due to confidentiality issues for participating students and teaching physicians but are available from the corresponding author on reasonable request.

## Oversight and monitoring

### Composition of the coordinating centre and trial steering committee {5d}

Not applicable, as this is a teaching quality improvement project.

### Composition of the data monitoring committee, its role and reporting structure {21a}

Not applicable, as this is a teaching quality improvement project.

### Adverse event reporting and harms {22}

Not applicable, as this is a teaching quality improvement project.

### Frequency and plans for auditing trial conduct {23}

Not applicable, as this is a teaching quality improvement project.

### Plans for communicating important protocol amendments to relevant parties (e.g. trial participants, ethical committees) {25}

In case of modifications of the protocol, the above-mentioned ethics committee will be informed.

## Dissemination plans {31a}

The study findings will be disseminated by journal publications and congress presentations.

## Discussion

To our knowledge, this is the first controlled trial that studies the effectiveness of a toolbox concept with EPAs for undergraduate training in family practices. We expect a significant improvement in students’ overall satisfaction with their learning progress by the intervention.

## Trial status

The intervention will start with the first education of teaching physicians.

## Supporting information

SPIRIT Checklist

Ethics Statement

Funding Documentation

## Data Availability

All data produced in the present study will be available upon reasonable request to the authors

## Abbreviations

EPA: Entrustable Professional Activity
TFM: Toolbox Family Medicine

## Declarations

## Acknowledgements

We owe thanks to the Medical Faculty of the University of Bonn for providing financial support for a student research assistant within the Program for Teaching Quality. In addition, we thank the team of teaching physicians for their support in developing and reviewing the EPAs.

## Authors’ contributions {31b}

BW conceived the study. NA drafted the first version of the manuscript together with BW and DW. NA, JS and MW developed the content of the toolbox index cards and their design. TW performed the sample size calculation and advised on the statistical approach. All authors contributed to the study protocol and the development of the intervention. All authors provided feedback on the manuscript and approved the final version.

## Availability of data and materials {29}

Only scientists of the Institute of General Practice and Family Medicine, University of Bonn, will have access to the final pseudonymized dataset. A master file with names of the participating teaching physicians will be stored in the access-restricted teaching databank of the institute.

## Ethics approval and consent to participate {24}

This study complies with the ethical principles of the World Medical Association Declaration of Helsinki. As the study constitutes a teaching quality improvement project without risks to the participants, the Ethics Committee of the Medical Faculty of the University of Bonn deemed it exempt from review on August 26, 2021. Participants are informed on the purpose and the voluntary nature of the evaluation. Participation in the online-evaluation is considered informed consent.

## Consent for publication {32}

Not applicable. No data relating to an individual person will be published.

## Authors’ information (optional)

Not applicable.

## References

Chen HC, van den Broek WES, Ten Cate O. The case for use of entrustable professional activities in undergraduate medical education. Academic medicine. 2015; doi: 10.1097/ACM.0000000000000586

Cooper S, Cant R, Waters D, et al. Measuring the quality of nursing clinical placements and the development of the Placement Evaluation Tool (PET) in a mixed methods co-design project. BMC nursing. 2020; doi: 10.1186/s12912-020-00491-1.

Happ M, Bathke AC, Brunner E. Optimal sample size planning for the Wilcoxon-Mann-Whitney test. Statistics in medicine. 2019; doi: 10.1002/sim.7983.

Herwig A, Viehmann A, Thielmann A, et al. Relevance of clerkship characteristics in changing students’ interest in family medicine: a questionnaire survey. BMJ open. 2017; doi: 10.1136/bmjopen-2016-012794.

Holzhausen Y, Maaz A, Renz A, et al. Development of Entrustable Professional Activities for entry into residency at the Charité Berlin. GMS journal for medical education. 2019; doi: 10.3205/zma001213.

Pershing S, Fuchs VR. Restructuring medical education to meet current and future health care needs. Academic Medicine. 2013; doi: 10.1097/ACM.0000000000000020.

Peters H, Holzhausen Y, Boscardin C, et al. Twelve tips for the implementation of EPAs for assessment and entrustment decisions. Medical teacher. 2017; doi: 10.1080/0142159X.2017.1331031.

Schick K, Eissner A, Wijnen-Meijer M, et al. Implementing a logbook on entrustable professional activities in the final year of undergraduate medical education in Germany - a multicentric pilot study. GMS journal for medical education. 2019; doi: 10.3205/zma001277.

Schroeder J, Wehner M, Amarell N, et al. Evaluation of practice rotations using the Placement Evaluation Tool (PET): A standardized translation involving medical students and teaching physicians. Poster presented at: 93rd EGPRN Meeting, October 16, 2021; Halle (Saale) Germany.

Ten Cate O, Chen HC, Hoff RG, et al. Curriculum development for the workplace using Entrustable Professional Activities (EPAs). Medical Teacher. 2015; doi: 10.3109/0142159X.2015.1060308.

Van Buuren S. Flexible Imputation of Missing Data. 2^nd^ ed. Chapman & Hall/CRC Interdisciplinary Statistics Series; 2018.

