## Supplementary material for "Effectiveness of the EPA-based ‘Toolbox Family Medicine’ on students’ learning satisfaction: study protocol for a controlled trial": Ethics Statement

**Translation of Ethics-Committee document (August 26, 2021)**

To:
Prof. Birgitta Weltermann
Chair, Institute of Family Medicine and General Practice
Venusberg-Campus 1
53127 Bonn

From:
Prof. Kurt Racké
Ethics Committee
Medical Faculty, University of Bonn
Venusberg-Campus 1
53127 Bonn

Date: August 26, 2021
KRa/AS

**Evaluation of General Practice Rotation (Blockpraktikum)**
**Your request via email from August 25, 2021**

Dear Mrs. Weltermann,

The above-mentioned documents were evaluated in the simplified process by the Chair of the Ethics' Committee of the Medical Faculty of Rheinische-Fredrich-Wilhelms-University Bonn:

It is confirmed that the described evaluation of the course "Blockpraktikum Allgemeinmedizin" (Family Medicine rotation) is not considered "biomedical research on humans" and thus this research project does not need counseling by the ethics committee in accordance with § 15 of the professional code for the physicians in North-Rhine.

Yours sincerely,

Prof. Dr. Kurt Racké
Chair Ethics Committee

**Contact Information**

Prof. Birgitta Weltermann
Institute of Family Medicine and General Practice
Venusberg-Campus 1
53127 Bonn, Germany

Prof. Dr. Birgitta Weltermann MPH(USA)
Direktorin, Institut für Hausarztmedizin
Haus 05, 1.OG Raum 270
Venusberg-Campus 1, 53127 Bonn
Tel. 0228/287-11156, Fax -11160
