## Supplementary material for "Effectiveness of the EPA-based ‘Toolbox Family Medicine’ on students’ learning satisfaction: study protocol for a controlled trial": Funding Documentation

**Your proposal to the quality improvement committee / Meeting June 30, 2021**

June 30, 2021

Dear Prof Weltermann,

You made a proposal to the above mentioned quality improvement committee meeting requesting funding for the intervention "Toolbox General Practice – student assistant".

We kindly inform you that the Dean's Office has agreed to support your project with 13,970.67 €.

[...]

Yours sincerely,

Prof. Dr. B. Pötsch
Vice Dean Teaching and Studies

Dean's office, Medical Faculty of the University of Bonn
Venusberg-Campus 1
53127 Bonn

**Contact Information**

Prof. Birgitta Weltermann
Institute of Family Medicine and General Practice
Venusberg-Campus 1
53127 Bonn, Germany

Prof. Dr. Birgitta Weltermann MPH(USA)
Direktorin, Institut für Hausarztmedizin
Haus 05, 1.OG Raum 270
Venusberg-Campus 1, 53127 Bonn
Tel. 0228/287-11156, Fax -11160
